# Ethnic inequalities among NHS staff in England - workplace experiences during the COVID-19 pandemic

**DOI:** 10.1101/2023.04.13.23288481

**Authors:** Rebecca Rhead, Lisa Harber-Aschan, Juliana Onwumere, Catherine Polling, Sarah Dorrington, Annahita Ehsan, Sharon AM Stevelink, Kamlesh Khunti, Ghazala Mir, Richard Morriss, Simon Wessely, Charlotte Woodhead, Stephani L Hatch

## Abstract

**Objectives:** To determine how workplace experiences of NHS staff varied by ethnic group during the COVID-19 pandemic and examine how these experiences are associated with mental and physical health at the time of the study.

**Methods:** An online Inequalities Survey was conducted by the TIDES study (Tackling Inequalities and Discrimination Experiences in health Services) in collaboration with NHS CHECK. This Inequalities Survey collected measures relating to workplace experiences (such as personal protective equipment (PPE), risk assessments, redeployments, and discrimination) as well as mental health, and physical health from NHS staff working in the 18 trusts participating with the NHS CHECK study between February and October 2021 (N=4622).

**Results:** Regression analysis revealed that staff from Black and Mixed/Other ethnic groups had greater odds of experiencing workplace harassment (adjusted odds ratio (AOR) = 2.43 [1.56-3.78] and 2.38 [1.12-5.07], respectively) and discrimination (AOR = 4.36 [2.73-6.96], and 3.94 [1.67-9.33], respectively) compared to White British staff. Staff from black ethnic groups also had greater odds than White British staff of reporting PPE unavailability (AOR = 2.16 [1.16-4.00]). Such workplace experiences were associated with negative physical and mental health outcomes, though this association varied by ethnicity. Conversely, understanding employment rights around redeployment, being informed about, and having the ability to inform redeployment decisions were associated with lower odds of poor health outcomes.

**Conclusions:** Structural changes to the way staff from ethnically minoritised groups are supported, and how their complaints are addressed by leaders within the NHS are urgently required to address racism and inequalities in the NHS.

**Policy implications:** Maintaining transparency and implementing effective mechanisms for addressing poor working conditions, harassment, and discrimination is crucial in the NHS. This can be achieved through appointing a designated staff member, establishing a tracking system, and training HR managers in identifying and handling reports of racial discrimination. Incorporating diversity and inclusion considerations into professional development activities and providing staff with opportunities to actively participate in decision-making can also benefit their health. The NHS Workforce Race Equality Standard may need to broaden its scope to assess race equality effectively.

## Introduction

Staff from ethnically minoritised groups constitute approximately 22% of the National Health Service (NHS) workforce in England (50% in London), but are underrepresented in senior roles, more likely to face disciplinary action, and experience less control over their working conditions compared to White staff ^1,2^. Furthermore, the NHS Workforce Race Equality Standard (WRES) – a programme designed to monitor race equality in the NHS – has consistently found that staff from ethnically minoritised groups experience disproportionate levels of discrimination and harassment from patients and colleagues (particularly the latter) ^3,4^. Such experiences negatively impact mental and physical health and are associated with long periods of sickness absence^5^. Qualitative research has also found that staff from ethnically minoritised groups working in London NHS Trusts may cope with bullying and microaggressions by moving teams or leaving their jobs ^2^.

Within the UK, healthcare staff from ethnic minorities have been over-represented in deaths due to COVID-19 ^6–11^. Reasons for this are complex but are partially the result of long-standing structural racism which has concentrated staff from ethnically minoritised groups in lower grades with more front-line work and greater exposure to COVID-19 ^3^. Recent commentaries suggest staff from ethnic minorities were more likely to be redeployed into hospital/clinic areas with a high risk of COVID-19 during the pandemic because they were unable to challenge or inform these decisions ^12^. Though COVID-19 risk assessments were introduced in April 2020 to ensure safe working conditions for all staff ^13^, these may have enabled further workplace inequalities if not conducted fairly.

Inequalities in COVID-19 exposure have also been compounded by disproportionately inadequate access to personal protective equipment (PPE). A survey of 1,119 ethnically diverse UK healthcare staff during the pandemic found that 96% of ethnically minoritised participants believed that inadequate PPE had directly contributed to the transmission of COVID-19 in healthcare staff (vs 75% of White participants)^14^. Ethnically minoritised respondents were more likely to report concerns about PPE and to feel unable to decline requests to work in the absence of adequate PPE. These findings were echoed in a UK-based survey of 4,418 nursing staff which found that staff from ethnic minorities were more likely than White British staff to report problems accessing PPE, feel pressured to provide care without it, and have unaddressed PPE concerns^15^. Similarly, UK-REACH (United Kingdom Research study into Ethnicity And COVID-19 outcomes in Healthcare staff) analysed data gathered between December 2020 and February 2021 from over 10,000 healthcare staff, finding that Asian ethnic staff groups were less likely to report access to adequate PPE compared to White staff groups^16^. Finally, a qualitative study of 53 NHS staff & leaders, service users and community partners from ethnically minoritised backgrounds interviewed during the pandemic found that staff feared speaking up about working conditions would affect future employment. This was particularly true for agency/temp staff and those whose immigration status increased their precarity^17^. Findings from these studies reflect the higher rates of COVID-19 and greater social risk factors for minority ethnic groups more widely.

Evidence suggests that pressurised working environments (e.g., high workload, short staffing) can exacerbate bullying and discrimination^2,18^. This may disproportionately impact staff from ethnically minoritised groups due to their overrepresentation at lower levels of the workforce hierarchy, negative stereotyping, and prevailing organisational norms ^2^. Therefore, the extraordinary pressures of the COVID-19 pandemic may have potentially increased rates of bullying, harassment, and discrimination for ethnically minoritised NHS staff, further impacting their mental and physical health. However, existing research examining this is either limited ^12^, or focuses on a single region or role ^14,15,19^.

NHS CHECK is one of the largest UK cohort studies conducted during the pandemic, established in April 2020 to longitudinally investigate the psychosocial impacts of the COVID-19 pandemic on NHS staff. The ongoing online survey assesses the mental health and well-being of NHS staff, students and volunteers within 18 NHS Trusts ^20^. Initial findings from NHS CHECK indicated that women, younger staff and nurses in London Trusts reported poorer mental health outcomes than other staff between April and June 2020 ^21^.

However, these analyses did not examine inequities by ethnicity.

Given the ongoing and evolving pressures beyond the COVID-19 pandemic, it is vital to recognise any persistent inequalities that may have led to negative health and job-related consequences for ethnically minoritised NHS staff. Thus, the Tackling Inequalities and Discrimination Experiences in health Services (TIDES) study partnered with NHS CHECK to develop a survey to capture inequalities during the COVID-19 pandemic (the TIDES Inequalities Survey). This paper aims to:

1. Estimate the prevalence of negative workplace experiences (e.g., PPE unavailability, bullying) during the pandemic by ethnic groups.
2. Examine to what extent such experiences were associated with physical and mental health outcomes.

## Methods

The TIDES study investigates ethnic health inequalities and discrimination in UK health and social care providers (www.tidesstudy.com). Together with NHS peer researchers (healthcare staff trained in research methods) and national advisory and stakeholder opinion groups, TIDES co-designed an Inequalities Survey to be incorporated into the 10-month follow-up of NHS CHECK study. This survey was designed to assess the impact of COVID-19 on ethnic inequalities experienced by NHS staff.

The Inequalities Survey was compiled through a modified Delphi consensus process ^22^ involving discussions and prioritisation surveys with frontline NHS staff, senior NHS managers/leaders and NHS equality, diversity and inclusion leaders about their experiences during the pandemic. The consensus-building approach was iterative, including several stakeholder discussions, piloting of proposed survey questions, and refining accordingly (Figure 1). During these discussions, staff described experiences of PPE, poorly performed COVID-19 workplace risk assessments, sudden re-deployments that could not be challenged or discussed, and experiences of discrimination and harassment in the workplace. All were described as disproportionately affecting ethnically minoritised staff. Questions on these topics and socioeconomic, occupational and COVID-19 related questions were included in the survey.

**Figure 1:**
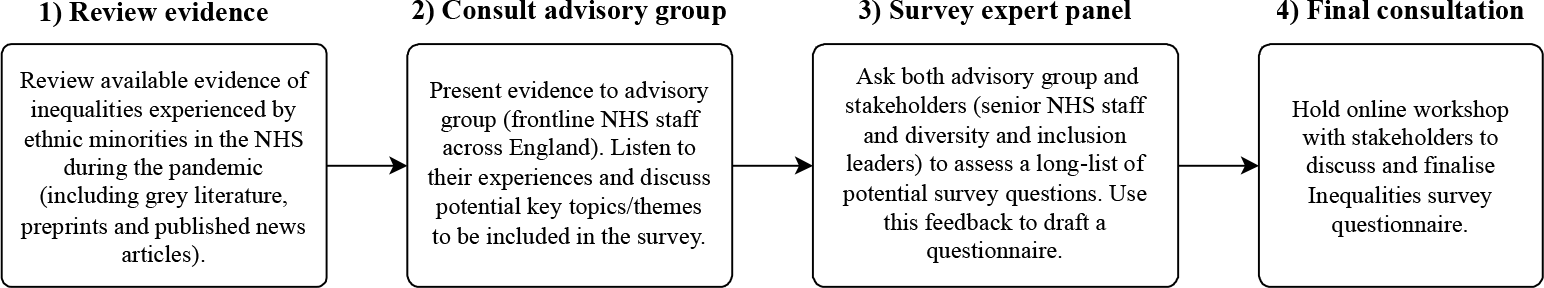
Consensus building process of working with stakeholders and advisory groups to produce the inequalities survey questionnaire.

### Data

NHS CHECK invited all NHS staff (including medical, nursing, midwifery, allied health professionals, support, administrative, management, volunteers, and students fast-tracked into clinical roles) working in 18 NHS Trusts across England (see Supplementary Material for the full list of Trusts) to participate in their study ^21^.

The baseline NHS CHECK survey comprised two versions: a mandatory and expedited version, as well as an optional, more detailed version. However, both versions were relatively brief in nature. By contrast, subsequent follow-up surveys were characterized by greater length and complexity, including the incorporation of supplementary measures that were not present in the initial survey. In most participating Trusts, there was strong support for the baseline survey to be completed from senior NHS management as well as e-mail and text reminders to staff. COVID restrictions prevented researchers from gaining face-to-face access to staff and many frontline staff could not access e-mail or personal phones in the workplace.

There were no monetary or other incentives to take part, but participants were entered into a prize draw (Lamb et al. (2021) describe further recruitment details). The NHS CHECK baseline sample consisted of 23,446 participants across 18 Trusts (total Trust population=139,037; response rate 5.9%) ^20^, followed by a 6-month follow-up (N=10,671; response rate 45.5%) and a 12-month follow-up (response rate unknown at the time of publication). All baseline participants were invited to participate in the Inequalities survey (10-month follow-up, N=4,622; response rate 19.7%) via email between February and October 2021 (Figure 2). Participation in the Inequalities Survey involved completing an online questionnaire ^23^.

**Figure 2:**
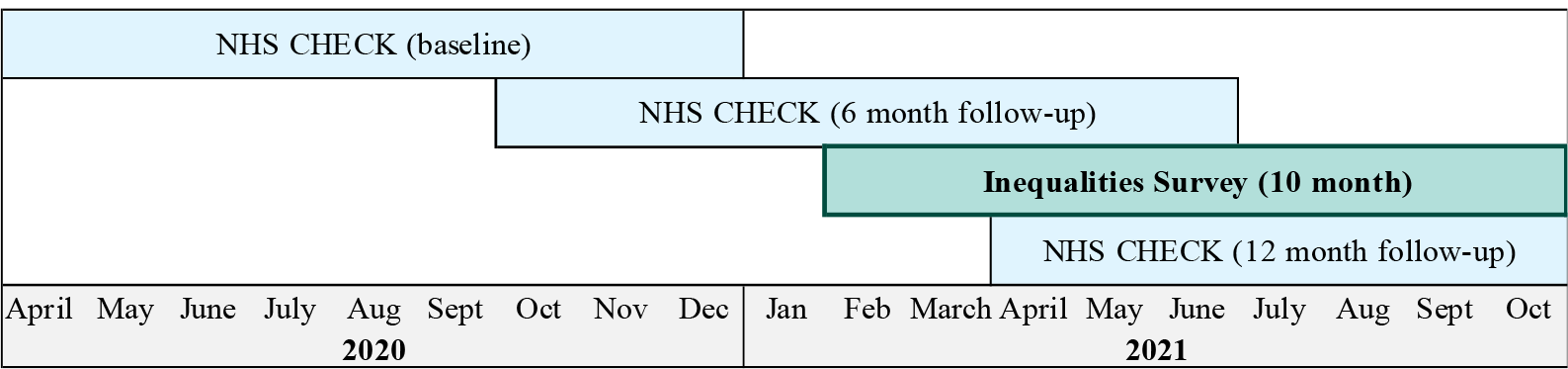
Timelines for the TIDES Inequalities Survey alongside NHS CHECK surveys

### Health outcomes

Measures to assess probable depression, anxiety and somatic symptoms were included in the survey. The Patient Health Questionnaire (PHQ-9) assessed depression, using a score of ≥10 to indicate probable depressive disorder (88% sensitivity and specificity for major depression ^24^). The GAD-7 scale assessed anxiety, using a score of ≥8 to indicate probable anxiety disorder (89% sensitivity, 82% specificity) ^25^. Both scales have very good to excellent levels of internal consistency and test-retest reliability ^26^. The PHQ-15 somatic symptom subscale assessed physical health ^27,28^, where 15 symptoms (e.g., pain, nausea, fatigue) are rated as 0 (“not bothered at all”), 1 (“bothered a little”), or 2 (“bothered a lot”), producing a score of 0-30. A score of ≥10 indicated moderate to severe somatic symptoms.

### Negative Workplace Experiences

Participants were asked if they had ever been unable to access PPE when at work during the pandemic and whether they had received a COVID-19 risk assessment. Participants were also asked if they had been redeployed during the pandemic and whether they had a good understanding of their employment rights relating to redeployment. If they had been redeployed, they were also asked if they were forewarned, or able to discuss or challenge the redeployment.

Discrimination was assessed by asking “In the last 12 months have you personally *experienced* discrimination at work from a manager/team leader or other colleagues”. The item assessing bullying, harassment and abuse (BHA) asked “In the last 12 months how many times have you personally *experienced* BHA from managers?” and “In the last 12 months how many times have you personally *experienced* BHA from other colleagues?”. These items were combined and dichotomised to produce a single measure of whether the participant had experienced discrimination or BHA from any co-worker. These measures of discrimination and BHA were taken from the NHS’s Staff Survey ^29^. See Supplementary Material for a full list of questions.

### Analysis

Descriptive statistics described the Inequalities Survey sample by:

- Age (≤30, 31-40, 41-50, ≥51)
- Sex (male, female)
- Migrant status (yes, no)
- Region (London, South, North)
- Job role (doctor, nurse, other clinical, non-clinical)
- Employment contract (permanent contract, bank/agency shifts, both)
- Ethnicity (defined by UK Census categories ^30^, collected at baseline). Due to small sample sizes for specific ethnic groups (n<10), ethnicity was aggregated into five categories: White British, White Other, Black, Asian and Mixed/Other (see Supplementary Material for details of the ethnic groups included in these categories)

The prevalence of workplace experiences was calculated overall and for each ethnic group. Logistic regression was used to examine associations between i) ethnicity and specific workplace experiences, and ii) workplace experiences and mental health outcomes (probable anxiety, probable depression, moderate/severe somatic symptoms). These regression analyses estimated unadjusted odds ratios (ORs), and ORs adjusted for age, sex, region, contract, job role and month of survey completion (decided *a priori* and informed by relevant literature ^2,5^). Additional subgroup analyses assessed the impact of BHA and discrimination experiences on probable depression for specific ethnic groups; sample size restrictions did not allow for subgroup analyses for other exposures and outcomes. Response weights were generated for the baseline NHS CHECK survey using iterative proportional fitting (a raking algorithm) based on age, gender, ethnicity, and role. To use these weights, Inequalities Survey participants were treated as a sub-population of the full (baseline) sample using the Survey packages’ subset command ^31^. This allowed our analysis to use the original design information from the baseline data but restrict survey design to participants of the Inequalities Survey. The alternative of dropping those who did not participate in the Inequalities Survey would produce correct estimates but incorrect standard errors ^32^. Finally, post-estimation commands from the Survey package were used to account for Trust size and response rate. Reported prevalence estimates, ORs and 95% confidence intervals (CI) were weighted, and frequencies were unweighted. All analysis was conducted in R V.4.2.0. ^33^ using the Survey package ^31^.

## Results

The demographic composition of the Inequalities Survey sample (n=4622) is similar to that of the baseline NHS CHECK survey. As shown in the Supplementary Material, gender composition is the same across both samples and Inequalities Survey participants are slightly older. The composition of ethnic groups is broadly similar across both samples though the Inequalities Survey has a slightly higher percentage of staff who belong to the White Other ethnic group (8.5% vs 6.3%) and slightly lower percentages of Black (2.9% vs 4.3%) and Asian groups (4.8% vs 6.6%).

As shown in Table 1, most of the sample were female (75%), born in the UK (84%), worked in clinical roles (68%), and had a permanent employment contract (90%). Almost half of the staff from Black ethnic groups worked in non-clinical roles (compared to 33% in the White British group) and over two-thirds were London-based (compared to 13% in White British group). In contrast, staff from Asian ethnic groups were predominantly employed in clinical roles and had the highest proportion of doctors (20%), almost half were based in Northern England. Staff from the Mixed/Other group (mostly represented by mixed White and Asian groups) had the highest proportion of nurses (33%) among all other ethnic groups. Staff from White Other groups had a similar composition of job roles as the White British group, but a higher proportion (82%) worked in the South/London region.

**Table 1:** Sociodemographic, work and health characteristics of Inequalities Survey participants by ethnicity

|  | **Overall**<br>**N = 4622** | **White British**<br>**n = 3741** | **White Other**<br>**n = 392** | **Black**<br>**n = 136** | **Asian**<br>**n = 220** | **Mixed/Other**<br>**n = 133** |
| --- | --- | --- | --- | --- | --- | --- |
| **Age (years) category** |  |  |  |  |  |  |
| ≤30 | 645 (15.0%) | 503 (14.4%) | 51 (11.6%) | 17 (12.4%) | 44 (22.1%) | 30 (26.1%) |
| 31-40 | 923 (22.8%) | 692 (21.0%) | 112 (32.5%) | 28 (21.1%) | 69 (33.9%) | 22 (14.2%) |
| 41-50 | 1,290 (25.6%) | 1,045 (25.5%) | 102 (22.3%) | 43 (28.7%) | 60 (25.6%) | 40 (36.0%) |
| 51+ | 1,764 (36.5%) | 1,501 (39.2%) | 127 (33.6%) | 48 (37.8%) | 47 (18.4%) | 41 (23.8%) |
| **Gender** |  |  |  |  |  |  |
| Female | 3,725 (75.3%) | 3,050 (76.1%) | 307 (69.8%) | 107 (76.0%) | 162 (73.6%) | 99 (68.6%) |
| Male | 825 (23.1%) | 640 (22.3%) | 77 (27.2%) | 25 (21.4%) | 53 (25.4%) | 30 (29.4%) |
| Other | 72 (1.7%) | 51 (1.6%) | 8 (3.0%) | 4 (2.5%) | 5 (1.0%) | 4 (2.0%) |
| **Migration** |  |  |  |  |  |  |
| UK born | 3,964 (83.8%) | 3,615 (96.9%) | 91 (25.8%) | 73 (51.2%) | 97 (45.0%) | 88 (54.5%) |
| Born outside UK | 632 (16.2%) | 105 (3.1%) | 299 (74.2%) | 61 (48.8%) | 122 (55.0%) | 45 (45.5%) |
| **Job role** |  |  |  |  |  |  |
| Doctor | 300 (9.3%) | 200 (8.1%) | 38 (11.9%) | 9 (8.3%) | 48 (20.0%) | 5 (3.7%) |
| Nurse | 1,073 (27.7%) | 868 (28.1%) | 84 (27.5%) | 38 (26.4%) | 50 (24.5%) | 33 (33.0%) |
| Other clinical | 1,263 (30.4%) | 1,018 (31.0%) | 121 (30.4%) | 31 (20.0%) | 49 (30.1%) | 44 (30.9%) |
| Non-clinical | 1,941 (32.5%) | 1,615 (32.8%) | 146 (30.1%) | 57 (45.3%) | 73 (25.3%) | 50 (32.4%) |
| **Contract** |  |  |  |  |  |  |
| Permanent only | 3,522 (74.4%) | 2,926 (76.6%) | 265 (68.0%) | 88 (68.3%) | 143 (66.4%) | 100 (65.5%) |
| Permanent with some bank shifts | 559 (15.7%) | 417 (14.3%) | 58 (18.0%) | 27 (18.2%) | 40 (21.6%) | 17 (26.1%) |
| Bank shifts only | 148 (3.0%) | 109 (2.8%) | 19 (4.5%) | 11 (6.7%) | 4 (2.0%) | 5 (1.0%) |
| Other | 352 (6.9%) | 257 (6.3%) | 45 (9.5%) | 9 (6.9%) | 30 (10.0%) | 11 (7.3%) |
| **Region** |  |  |  |  |  |  |
| London | 1,065 (20.5%) | 638 (13.4%) | 196 (42.3%) | 89 (71.3%) | 92 (29.4%) | 50 (41.3%) |
| South | 1,951 (41.5%) | 1,697 (45.9%) | 134 (39.6%) | 16 (11.6%) | 54 (24.4%) | 50 (29.1%) |
| North | 1,606 (38.0%) | 1,406 (40.7%) | 62 (18.1%) | 31 (17.2%) | 74 (46.2%) | 33 (29.7%) |
| **Unavailable PPE (if applicable)** |  |  |  |  |  |  |
| No | 2,756 (81.8%) | 2,260 (82.1%) | 220 (78.6%) | 69 (70.4%) | 126 (90.0%) | 81 (76.3%) |
| Yes | 570 (18.2%) | 450 (17.9%) | 55 (21.4%) | 27 (29.6%) | 23 (10.0%) | 15 (23.7%) |
| **Risk assessment received** |  |  |  |  |  |  |
| No | 931 (23.3%) | 777 (24.3%) | 94 (24.4%) | 13 (11.2%) | 24 (17.1%) | 23 (33.5%) |
| Yes | 2,898 (76.7%) | 2,340 (75.7%) | 231 (75.6%) | 96 (88.8%) | 143 (82.9%) | 88 (66.5%) |
| **Redeployed to another role** |  |  |  |  |  |  |
| No | 2,703 (65.4%) | 2,228 (65.5%) | 213 (65.5%) | 75 (65.9%) | 111 (67.4%) | 76 (54.1%) |
| Yes | 1,123 (34.6%) | 886 (34.5%) | 112 (34.5%) | 34 (34.1%) | 56 (32.6%) | 35 (45.9%) |
| **Experienced BHA from staff** |  |  |  |  |  |  |
| No | 2,738 (66.0%) | 2,252 (67.4%) | 229 (64.3%) | 65 (46.7%) | 124 (70.8%) | 68 (47.4%) |
| Yes | 1,253 (34.0%) | 981 (32.6%) | 111 (35.7%) | 53 (53.3%) | 59 (29.2%) | 49 (52.6%) |
| **Experienced discrimination from staff** |  |  |  |  |  |  |
| No | 3,254 (80.0%) | 2,711 (83.3%) | 260 (74.6%) | 68 (52.4%) | 136 (77.1%) | 79 (55.6%) |
| Yes | 737 (20.0%) | 522 (16.7%) | 80 (25.4%) | 50 (47.6%) | 47 (22.9%) | 38 (44.4%) |
| **Probable depression (PHQ-9 score ≥10)** |  |  |  |  |  |  |
| No | 3,015 (77.0%) | 2,469 (77.8%) | 245 (73.1%) | 85 (75.4%) | 128 (78.0%) | 88 (63.6%) |
| Yes | 871 (23.0%) | 687 (22.2%) | 83 (26.9%) | 28 (24.6%) | 46 (22.0%) | 27 (36.4%) |
| **Probable anxiety (GAD-7 score ≥8)** |  |  |  |  |  |  |
| No | 3,244 (82.4%) | 2,660 (83.3%) | 264 (80.0%) | 96 (86.8%) | 129 (76.1%) | 95 (72.0%) |
| Yes | 642 (17.6%) | 496 (16.7%) | 64 (20.0%) | 17 (13.2%) | 45 (23.9%) | 20 (28.0%) |
| **Moderate/severe somatic symptoms (PHQ-15 score ≥10)** |  |  |  |  |  |  |
| No | 2,947 (76.6%) | 2,415 (77.0%) | 235 (69.6%) | 88 (76.9%) | 131 (81.5%) | 78 (66.7%) |
| Yes | 922 (23.4%) | 729 (23.0%) | 92 (30.4%) | 25 (23.1%) | 40 (18.5%) | 36 (33.3%) |

Total cell counts may vary due to missing data.
PPE = Personal Protective Equipment
BHA = Bullying, Harassment or Abuse
Bank/agency shifts are temporary shifts at trust hospitals to cover staff absences.
See Supplementary Material for full list of trusts per region

Across the sample, 23% indicated probable depression, 18% indicated probable anxiety, and 23% reported medium/severe somatic symptoms. Staff from the Mixed/Other ethnic group had a higher prevalence of probable depression, anxiety, and somatic symptoms than all other ethnic groups (36%, 28%, and 33% respectively). One third and one fifth of all survey respondents reported experiences of BHA and discrimination respectively (see Table 1).

### Prevalence of workplace experiences by ethnicity

In Table 2, findings indicate that staff from the Black ethnic group had greater odds of receiving a risk assessment (adjusted OR = 4.68, CI = 2.41,9.15) compared to staff from the White British group (see Table 2). However, they also had greater odds of reporting PPE unavailability (adjusted OR = 2.16, CI = 1.16,4.00). In contrast, staff from the Asian ethnic group had lower odds of reporting PPE unavailability (adjusted OR = 0.38, CI = 0.20,0.72) compared to staff from the White British group. Staff from both Black and Mixed/Other groups had greater odds of experiencing BHA (Black: adjusted OR = 2.43, CI = 1.56, 3.78], Mixed/Other: adjusted OR = 2.38, CI = 1.12, 5.07) as well as discrimination from other staff members (Black: adjusted OR = 4.36, CI=2.73, 6.96, Mixed/Other: adjusted OR = 3.94, CI=1.67, 9.33) compared to the White British group. Staff of White Other ethnicity also had greater odds of experiencing discrimination (adjusted OR = 1.61, CI=1.10, 2.35) compared to the White British group.

**Table 2:** Regression analysis to show associations between ethnicity and reported workplace experiences

|  | Unavailable PPE |  | Received risk assessment |  | Experienced BHA from staff |  | Experienced discrimination from staff |  | Redeployed |  |
| --- | --- | --- | --- | --- | --- | --- | --- | --- | --- | --- |
| Ethnicity | Crude OR [95%CI] | Adjusted OR [95%CI] | Crude OR [95%CI] | Adjusted OR [95%CI] | Crude OR [95%CI] | Adjusted OR [95%CI] | Crude OR [95%CI] | Adjusted OR [95%CI] | Crude OR [95%CI] | Adjusted OR [95%CI] |
| White Other | 1.24<br>[0.83, 1.87] | 1.04<br>[0.69, 1.56] | 1.00<br>[0.74, 1.34] | 1.31<br>[0.96, 1.78] | 1.15<br>[0.84, 1.56] | 1.14<br>[0.83, 1.56] | 1.70<br>[1.19, 2.42] | 1.61<br>[1.10, 2.35] | 1.00<br>[0.74, 1.35] | 0.85<br>[0.61, 1.17] |
| Black | 1.92<br>[1.14, 3.24] | 2.16<br>[1.16, 4.00] | 2.54<br>[1.33, 4.84] | 4.68<br>[2.41, 9.11] | 2.36<br>[1.57, 3.57] | 2.43<br>[1.56, 3.78] | 4.53<br>[2.96, 6.93] | 4.36<br>[2.73, 6.96] | 0.98<br>[0.62, 1.55] | 0.85<br>[0.52, 1.36] |
| Asian | 0.51<br>[0.27, 0.96] | 0.38<br>[0.20, 0.72] | 1.56<br>[0.73, 3.32] | 2.21<br>[1.01, 4.84] | 0.86<br>[0.54, 1.36] | 0.84<br>[0.52, 1.33] | 1.48<br>[0.91, 2.42] | 1.53<br>[0.93, 2.52] | 0.92<br>[0.57, 1.48] | 0.74<br>[0.46, 1.19] |
| Mixed/Other | 1.42<br>[0.32, 6.31] | 1.22<br>[0.41, 3.70] | 0.64<br>[0.22, 1.82] | 0.86<br>[0.36, 2.10] | 2.29<br>[1.06, 4.99] | 2.38<br>[1.12, 5.07] | 3.99<br>[1.70, 9.37] | 3.94<br>[1.67, 9.33] | 1.61<br>[0.68, 3.84] | 1.31<br>[0.61, 2.79] |

*Adjusted models adjust for age, sex, region, contract, job role and month of survey completion.*
*White British is treated as the reference category.*
*BHA = Bullying, Harassment or Abuse*

### Redeployment decision

35% (n=1,123) of participants reported being redeployed into a different role during the pandemic (Table 1). Of those who were deployed, staff from the Black ethnic group had lower odds of feeling able to input into their redeployment (adjusted OR = 0.58, CI = 0.28, 1.20), while staff from the Mixed/Other group were less likely to be forewarned about their redeployment (adjusted OR = 0.23, CI = 0.09, 0.58, see Table 3). Staff from the Asian ethnic group had greater odds of feeling able to challenge their redeployment decision (adjusted OR = 3.17, CI = 1.26, 7.99). Of all participants (regardless of whether they were redeployed or not), staff from the Black ethnic group (35%, adjusted OR = 0.53, CI = 0.32, 0.86) were the only group less likely to understand their redeployment rights than White British staff (46% - see Supplementary Material).

**Table 3:** Regression analysis to show associations between ethnicity and redeployment experiences in those who were redeployed (n=1,123)

| Ethnicity | Able to challenge redeployment |  |  | Warned about redeployment |  |  | Able to have a say (input) about redeployment |  |  |
| --- | --- | --- | --- | --- | --- | --- | --- | --- | --- |
|  | n (%) | Crude OR<br>[95%CI] | Adjusted OR<br>[95%CI] | n (%) | Crude OR<br>[95%CI] | Adjusted OR<br>[95%CI] | n (%) | Crude OR<br>[95%CI] | Adjusted OR<br>[95%CI] |
| White British | 508 (50.6) | — | — | 650 (70.3) | — | — | 489 (51.1) | — | — |
| White Other | 66 (56.9) | 1.29<br>[0.81, 2.06] | 1.07<br>[0.65, 1.76] | 76 (67.3) | 0.87<br>[0.53, 1.43] | 0.74<br>[0.42, 1.30] | 59 (49.4) | 0.93<br>[0.59, 1.49] | 0.70<br>[0.44, 1.12] |
| Black | 15 (42.5) | 0.72<br>[0.34, 1.52] | 0.58<br>[0.28, 1.20] | 20 (62.1) | 0.69<br>[0.32, 1.49] | 0.68<br>[0.31, 1.53] | 12 (31.5) | 0.44<br>[0.20, 0.95] | 0.33<br>[0.15, 0.72] |
| Asian | 35 (72.4) | 2.56<br>[1.29, 5.08] | 3.17<br>[1.26, 7.99] | 41 (75.3) | 1.29<br>[0.60, 2.74] | 1.46<br>[0.59, 3.62] | 35 (68.5) | 2.08<br>[1.03, 4.20] | 2.38<br>[0.95, 5.97] |
| Mixed/Other | 15 (23.2) | 0.30<br>[0.09, 0.95] | 0.37<br>[0.14, 0.94] | 21 (32.2) | 0.20<br>[0.06, 0.68] | 0.23<br>[0.09, 0.58] | 18 (30.6) | 0.42<br>[0.13, 1.41] | 0.52<br>[0.21, 1.33] |

*Adjusted models adjust for age, sex, region, contract, job role and month of survey completion*

### Health outcomes by experience

As shown in Table 4, unavailable PPE was associated with an approximately twofold increase in probable depression, probable anxiety, and moderate/severe somatic symptoms. BHA and discrimination were also associated with an approximately three-fold increase in each of these health outcomes. Conversely, understanding redeployment rights was associated with lower odds of probable depression and moderate/severe somatic symptoms.

**Table 4:** Regression analysis to show associations between workplace experiences and mental and physical health outcomes

| Workplace Experience |  | Probable Depression<br>(PHQ-9 score ≥10) |  |  | Probable Anxiety<br>(GAD-7 score ≥8) |  |  | Moderate/Severe Somatic Symptoms<br>(PHQ-15 score ≥10) |  |  |
| --- | --- | --- | --- | --- | --- | --- | --- | --- | --- | --- |
|  |  | n (%) | Crude OR<br>[95%CI] | Adjusted OR<br>[95%CI] | n (%) | Crude OR<br>[95%CI] | Adjusted OR<br>[95%CI] | n (%) | Crude OR<br>[95%CI] | Adjusted OR<br>[95%CI] |
| Unavailable PPE | Yes | 192 (35.9) | 2.15<br>[1.62, 2.87] | 2.01<br>[1.52, 2.66] | 136 (27.5) | 2.01<br>[1.44, 2.80] | 1.73<br>[1.26, 2.36] | 1,380 (34.0) | 1.90<br>[1.42, 2.54] | 1.90<br>[1.43, 2.54] |
|  | No | 568 (20.6) | — | — | 428 (15.9) | — | — | 3,907 (21.0) | — | — |
| Risk assessment | Yes | 627 (22.2) | 0.81<br>[0.63, 1.04] | 0.86<br>[0.67, 1.10] | 450 (16.8) | 0.80<br>[0.60, 1.06] | 0.82<br>[0.62, 1.08] | 674 (22.4) | 0.77<br>[0.59, 0.99] | 0.80<br>[0.62, 1.04] |
|  | No | 236 (26.0) | — | — | 184 (20.2) | — | — | 242 (27.3) | — | — |
| Discrimination | Yes | 304 (43.8) | 3.59<br>[2.79, 4.62] | 3.65<br>[2.83, 4.70] | 226 (34.8) | 3.45<br>[2.61, 4.56] | 3.67<br>[2.79, 4.83] | 309 (41.8) | 3.10<br>[2.42, 3.98] | 2.99<br>[2.33, 3.85] |
|  | No | 567 (17.8) | — | — | 416 (13.4) | — | — | 613 (18.8) | — | — |
| BHA | Yes | 436 (35.7) | 2.84<br>[2.26, 3.55] | 3.02<br>[2.42, 3.77] | 341 (28.7) | 2.97<br>[2.31, 3.81] | 3.31<br>[2.58, 4.25] | 461 (36.5) | 2.88<br>[2.31, 3.59] | 3.00<br>[2.40, 3.75] |
|  | No | 435 (16.4) | — | — | 301 (11.9) | — | — | 461 (16.6) | — | — |
| Redeployed | Yes | 255 (23.4) | 1.02<br>[0.80, 1.30] | 0.97<br>[0.76, 1.24] | 190 (18.4) | 1.09<br>[0.82, 1.43] | 1.00<br>[0.76, 1.32] | 260 (24.0) | 1.05<br>[0.82, 1.33] | 0.98<br>[0.76, 1.26] |
|  | No | 607 (22.9) | — | — | 444 (17.2) | — | — | 655 (23.2) | — | — |
| Understand redeployment rights | Yes | 320 (18.5) | 0.62<br>[0.50, 0.77] | 0.66<br>[0.53, 0.83] | 232 (14.6) | 0.68<br>[0.53, 0.88] | 0.77<br>[0.60, 1.00] | 350 (18.3) | 0.59<br>[0.47, 0.73] | 0.62<br>[0.50, 0.77] |
|  | No | 541 (26.8) | — | — | 402 (20.0) | — | — | 561 (27.6) | — | — |
| Able to challenge redeployment<br>(if redeployed) | Yes | 121 (19.2) | 0.62<br>[0.42, 0.92] | 0.70<br>[0.48, 1.04] | 83 (14.3) | 0.57<br>[0.35, 0.91] | 0.68<br>[0.42, 1.12] | 114 (18.5) | 0.53<br>[0.35, 0.80] | 0.61<br>[0.41, 0.91] |
|  | No | 134 (27.8) | — | — | 107 (22.7) | — | — | 146 (29.9) | — | — |
| Warned about redeployment (if<br>redeployed) | Yes | 156 (18.9) | 0.47<br>[0.30, 0.73] | 0.53<br>[0.35, 0.79] | 119 (15.7) | 0.58<br>[0.35, 0.97] | 0.72<br>[0.44, 1.17] | 158 (20.1) | 0.51<br>[0.33, 0.80] | 0.58<br>[0.38, 0.88] |
|  | No | 99 (33.2) | — | — | 71 (24.3) | — | — | 102 (32.9) | — | — |
| Able to have a say (input) about<br>redeployment (if redeployed) | Yes | 122 (18.3) | 0.56<br>[0.38, 0.83] | 0.64<br>[0.43, 0.95] | 87 (14.4) | 0.58<br>[0.36, 0.93] | 0.70<br>[0.43, 1.16] | 117 (17.9) | 0.50<br>[0.33, 0.75] | 0.56<br>[0.38, 0.83] |
|  | No | 133 (28.6) | — | — | 103 (22.5) | — | — | 143 (30.4) | — | — |

*Adjusted models adjust for age, sex, region, contract, job role and month of survey completion*
*PPE = Personal Protective Equipment*
*BHA = Bullying, Harassment or Abuse*

Among those who were redeployed during the pandemic, having input into redeployment decisions, and being forewarned about redeployment were associated with lower odds of probable depression and moderate/severe somatic symptoms. Being able to challenge redeployment decisions was also associated with lower odds of experiencing moderate/severe somatic symptoms.

### Subgroup analysis

Descriptive subgroup analysis of probable depression by ethnicity, stratified by BHA and discrimination, indicated that across all ethnic groups, probable depression was more prevalent among those who reported these negative experiences, compared to those who did not (see Supplementary Material). However, the impact of these experiences on depression prevalence varied by ethnicity. Among those who did not experience BHA or discrimination at work, staff from the White British, Asian, and Black ethnic groups had the highest prevalence of depressive disorder. In contrast, among those who reported experiencing these negative experiences, staff from the White Other and Mixed/Other ethnic groups had the highest prevalence of depressive disorder. However, due to small sample sizes these finding cannot be generalised to the wider population.

## Discussion

Building on our pre-pandemic investigation into discrimination and inequalities in healthcare, this study aimed to identify ethnic inequalities in workplace experiences among NHS staff in England working during the COVID-19 pandemic. This work represents a collaborative effort with NHS CHECK and the NHS staff and leaders who comprised our advisory and stakeholder groups to inform the contents of our Inequalities Survey. Overall, this study found that negative workplace experiences such as discrimination, bullying and unavailable PPE were more likely to occur for staff from ethnically minoritised groups (particularly staff from Black and Mixed/Other ethnic groups). These workplace experiences were associated with negative physical and mental health outcomes. Conversely, understanding employment rights around redeployment, being warned about an upcoming redeployment, and being able to inform redeployment decisions were associated with lower odds of poor health outcomes.

The study found that the difference in the likelihood of experiencing probable depression among those who faced bullying, harassment, and discrimination varied by ethnicity. Specifically, the highest prevalence of depression was observed among the White Other and Mixed/Other groups who had experienced discrimination and BHA. Conversely, among those who did not experience BHA, staff from the Black and Mixed/Other ethnic groups had the highest prevalence of depression. However, these findings are based on small sample sizes, and thus, cannot be generalised. This highlights a broader issue where surveys often lack sufficient representation of ethnically minoritised groups to conduct effective subgroup analyses. To address this issue, academic researchers should prioritise building trust with communities to encourage participation in future studies. This can be achieved by involving community members and leaders in the survey design process, addressing concerns and offering incentives, providing transparency about the survey’s purpose and goals, partnering with trusted organisations, and ensuring ethical standards are upheld throughout the survey process. Such actions can help researchers to build relationships with communities, demonstrate a commitment to their concerns and interests, and contribute to a more inclusive and equitable research process.

Experiences of BHA and discrimination from staff were highly prevalent in our study and substantially higher among staff from all ethnically minoritised groups compared to estimates in the 2022 NHS Staff Survey ^1^. The overrepresentation of London Trusts in the Inequalities Survey data may have contributed to these higher prevalence estimates, as London Trusts have been known to perform poorly on these measures ^1,5^. The external nature of the Inequalities Survey might have encouraged greater disclosure of experiences of workplace discrimination and BHA. This was the case for the pre-pandemic TIDES survey which found higher rates of BHA and discrimination compared to the NHS Staff Survey ^5^.

Our research findings are consistent with previous studies, including the UK-REACH, which identified disparities in personal protective equipment (PPE) availability across different ethnic groups during the pandemic ^16^. Our study further contributes to the literature by demonstrating that inadequate PPE availability is associated with negative health outcomes among healthcare workers. This underscores the critical importance of ensuring equitable access to PPE (as well as a safe working environment for healthcare workers) during public health crises, particularly for healthcare workers from ethnically minoritised groups who are already vulnerable to health and socio-economic inequities. Our findings underscore the urgent need for evidence-based policies and interventions that prioritize equitable distribution of PPE to all healthcare workers, irrespective of their demographic characteristics, to promote health and safety during public health emergencies.

Our study also found alarmingly high exposure to negative workplace experiences related to harassment and discrimination among ethnically minoritised NHS staff during the pandemic. These findings are consistent with the most recent NHS staff survey ^1^ and UK-REACH ^34^ in addition to being supported by multiple qualitative studies that have explored similar workplace experiences among ethnically and racially minoritised groups ^35–37^. The short-term and long-term impacts of such experiences are likely to take a toll on the mental and physical health of employees ^38–40^, as well as their dependents and social networks, with implications for career progression, intention to remain at the NHS, and salary ^2^.

### Strengths and limitations

The Inequalities Survey represents one of the largest surveys examining the impact of the pandemic on inequalities among healthcare staff. Despite targeted efforts to increase engagement, the response rate from those who participated in the NHS CHECK baseline study was low and varied by ethnicity. Specifically, the response rate by ethnicity was 21% for White British staff, 27% for staff from White Other groups, 15% for staff from Black ethnic groups, 15% for staff from Asian ethnic groups, and 17% for staff from Mixed/Other ethnic groups. As a result, the relatively small sample sizes of staff from ethnically minoritised groups hindered our ability to examine the experiences of specific ethnic groups, such as Black Caribbean nurses. Additionally, conducting a thorough subgroup analysis to estimate the mental health impact of workplace experiences on specific ethnic groups was hampered by the same issue of limited sample sizes.

These sample size issues are partly due to recruitment being limited to participants from the NHS CHECK baseline survey, which had an overrepresentation of NHS staff from White ethnic groups (NHS CHECK = 86%, NHS workforce = 78% ^41^). Survey fatigue may also have contributed. Poor response rates from ethnically minoritised groups reflect a wider issue with UK public health surveys which typically include a disproportionately large proportion of participants from White ethnic groups ^42^. As highlighted in a recent Wellcome report, ethnically minoritised groups have demonstrated greater levels of mistrust in research and health institutions during the pandemic ^43^. The overrepresentation of ethnically minoritised staff at lower professional grades could also impact their ability to complete the survey if they have less control over their working patterns. Ideally, to overcome this in future studies, staff from lower grades should be given protected paid time off for research participation. This would increase participation rates and improve the representation of underrepresented groups in research studies.

Furthermore, the pandemic presented a particularly challenging context to recruit healthcare staff to research, given the stress experienced especially by ethnically minoritised staff. Nevertheless, a key strength of this survey was its tailored design to capture the unique experiences of ethnically minoritised NHS staff during these exceptional circumstances, by engaging staff and stakeholders through a consensus-building approach. To improve representation. In addition, the data were weighted based on age, gender, ethnicity, and role, using marginal socio-demographic data provided by participating trusts to ensure the sample better reflected our study population.

### Public Health Implications

The findings of this study provide additional evidence of the well-established link between racism, structural inequalities, and adverse health outcomes. It is crucial to prioritise racial discrimination as a public health issue, not just an ethical imperative, and ensure that decision-makers from ethically minoritised groups are involved in processes that affect their health and wellbeing. This requires the acknowledgement of the systemic nature of racism, as well as the implementation of robust systems to combat its key mechanisms, such as racial discrimination, among ethnically minoritised staff.

First, NHS Trusts must appoint designated staff who can work towards resolving reports of poor working conditions, harassment, and discrimination. Such posts should exist within each Trust and work as part of a network to institutional tackle discrimination, rather than as isolated appointments. Managers must also be trained to identify and handle reports of racial discrimination, with a shift in focus from generic cultural awareness and equality and diversity training, which has been found ineffective in tackling discrimination ^44,45^.

Alternative approaches such as interactive or experiential training ^46^ and inclusive leadership training ^47^, have been found to be more effective in addressing discrimination in the workplace. These approaches should also be incorporated into other professional development activities, such as leadership development programs, onboarding processes, and performance review systems.

Finally, this study identified health benefits for staff who understand their employment rights and are afforded opportunities to actively participate in decisions impacting their work. Consequently, NHS staff should be educated on their employment rights to ensure that they are able to advocate for themselves while also provided with adequate opportunities to engage in discussions, provide feedback, and question decisions concerning their working conditions. To effectively facilitate and monitor progress towards these goals in a transparent manner, the NHS Workforce Race Equality Standard may need to broaden its scope to include parameters such as tracking mechanisms for diversity and inclusion, as well as staff education initiatives. This would ensure that the NHS is actively monitoring and taking measures to improve in identified areas, while also ensuring that staff are equipped with the necessary knowledge and resources to create a more inclusive work environment. It would also aid in holding NHS leadership to account for addressing issues connected to diversity and inclusivity within their respective organisations.

## Conclusion

Against a backdrop of significant and publicised examples of health inequalities, discrimination, and economic instability, NHS staff have navigated challenging working environments throughout the COVID–19 pandemic. Our findings suggest that staff from ethnically minoritised groups have also been exposed to greater harassment, discrimination, and PPE unavailability than White British staff within the NHS, adding further burden to excess infection, mortality and need for intensive care among ethnic minorities ^10^. Indeed, given the high number of key worker status staff within the NHS and their responsibility for providing healthcare, findings strongly suggest that NHS staff should be afforded greater protection and support throughout the pandemic and beyond.

Addressing these problems requires structural transformation in terms of how staff from ethnically minoritised groups are supported and how their complaints are addressed, including urgent policy attention and mandatory representation in institutional decision-making ^48^. Additionally, educating staff on their employment rights is crucial to ensure that they are aware of their rights and are able to advocate for themselves. These approaches are urgently required to address racism and inequalities in the UK healthcare system, which have long been recognised as both ‘avoidable and unjust’^49^.

## Data Availability

All data produced in the present study are available upon reasonable request to the authors.

## Acknowledgements

The TIDES study team would like to acknowledge the invaluable contribution of our peer researchers, stakeholder opinion group, advisory group, collaborators and co-investigators. Without the continued support and expert knowledge of this group, this research would not have been possible.

