## Supplementary Material for "Ethnic inequalities among NHS staff in England - workplace experiences during the COVID-19 pandemic"

Ethnic inequalities experienced by NHS staff in England during the COVID-19 pandemic: Supplementary Material

### Supplementary Material A: Sample composition comparison

|  | **Baseline**   **(N = 22,834)** | **Inequalities Survey   (N = 4,622)** |
| --- | --- | --- |
| **Ethnicity** |  |  |
| White British | 18,127 (79%) | 3,741 (81%) |
| White Other | 1,446 (6%) | 392 (9%) |
| Black | 991 (4%) | 136 (3%) |
| Asian | 1,503 (7%) | 220 (5%) |
| Mixed/Other | 767 (3%) | 133 (3%) |
| **Gender** |  |  |
| female | 18,487 (81%) | 3,725 (81%) |
| male | 4,232 (19%) | 825 (18%) |
| other | 109 (1%) | 72 (2%) |
| **Age group (years)** | | |
| ≤30 | 4,367 (20%) | 645 (14%) |
| 31-40 | 5,068 (23%) | 923 (20%) |
| 41-50 | 5,794 (26%) | 1,290 (28%) |
| 51+ | 6,850 (31%) | 1,764 (38%) |
| Unweighted % | | |

### Supplementary Material B: Inequalities survey questionnaire

**Introduction**

The purpose of this follow-up survey is to understand the drivers of racial and ethnic inequalities in mental health and work outcomes particularly since COVID-19. This survey is for **ALL** staff, students and volunteers, not just those from ethnic minority backgrounds. It should take 15-20 minutes to complete.
  
This questionnaire has been designed by the [Tackling Inequalities and Discrimination Experiences in health Services (TIDES)](http://tidesstudy.com) study in collaboration with [NHS CHECK](http://nhscheck.org). 
  
Your responses will be used to identify racial and ethnic inequalities among staff and understand how COVID-19 might exacerbate such inequalities. 
  
This study is completely independent of any individual NHS trust or healthcare provider and is being carried out to develop training resources for those in managerial positions and inform NHS policies. 
  
Alongside our partners (Royal College of Nursing, Maudsley learning, NHS Confederation, Workforce Race Equality Standard, NHS England), we are working hard to ensure our research translates into real, positive impacts for NHS and social care staff from all racial and ethnic backgrounds.
  
Please read the [Information Sheet](https://kcliop.eu.qualtrics.com/CP/File.php?F=F_9pfmpYKv4XZKIYZ) for further information and contact details.

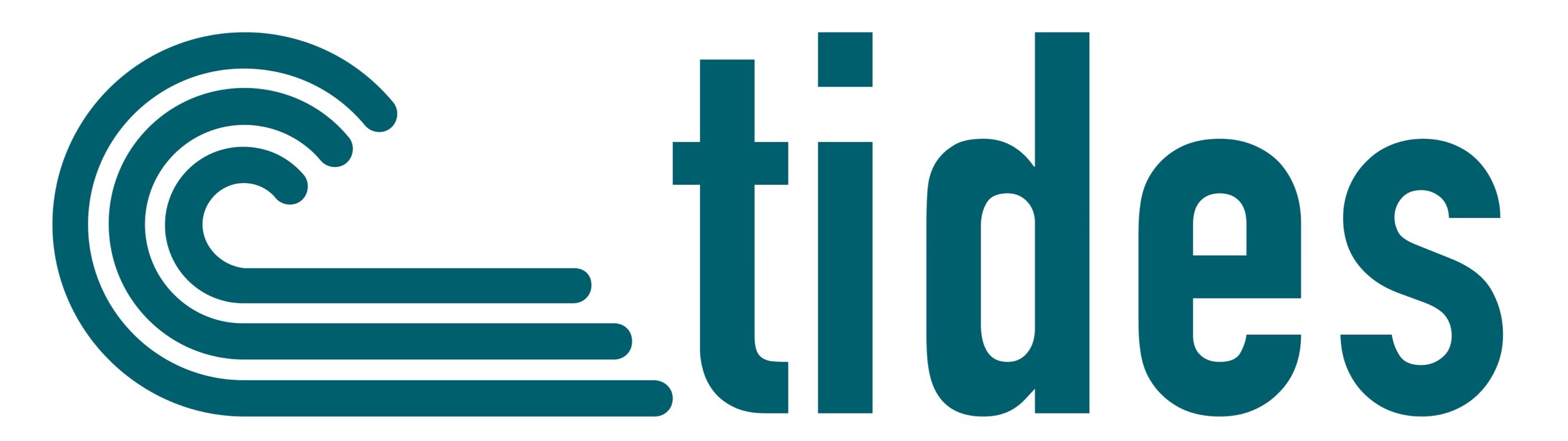

SECTION 1) Sociodemographic information

| **Name** | **Question** | **No** | **Option** | **Skip** |
| --- | --- | --- | --- | --- |
| image | What is your age? |  |  |  |
| Imgender1 | Which of the following best describes how you think of yourself? |  |  |  |
|  |  | 1 | Male (including trans man) | Skip to imgender2 |
|  |  | 2 | Female (including trans woman) | Skip to imgender2 |
|  |  | 3 | Non-binary | Skip to imgender2 |
|  |  | 4 | In another way |  |
|  |  | 5 | Prefer not to say | Skip to imgender2 |
| Imgender1_oth | Please specify |  |  |  |
| Imgender2 | Is this the same gender you were assigned at birth? |  |  |  |
|  |  | 1 | Yes |  |
|  |  | 2 | No |  |
|  |  | 3 | Prefer not to say |  |
| imorient | Do you identify as: |  |  |  |
|  |  | 1 | Heterosexual or straight |  |
|  |  | 2 | Lesbian |  |
|  |  | 3 | Gay |  |
|  |  | 4 | Bisexual |  |
|  |  | 5 | Other (e.g. Pansexual, Queer, Asexual, Unsure or questioning) |  |
|  |  | 6 | Prefer not to say |  |
| imrelig | What is your religion? |  |  |  |
|  |  | 1 | No religion | Skip to imlive |
|  |  | 2 | Buddhist | Skip to imlive |
|  |  | 3 | Christian (including Church of England, Catholic, Protestant & all other Christian denominations) | Skip to imlive |
|  |  | 4 | Hindu | Skip to imlive |
|  |  | 5 | Jewish | Skip to imlive |
|  |  | 6 | Muslim | Skip to imlive |
|  |  | 7 | Sikh | Skip to imlive |
|  |  | 8 | Other |  |
| Imrelig_oth | Please specify your religion: |  |  |  |
| imlive | Where do you live? |  |  |  |
|  |  | 1 | Urban area |  |
|  |  | 2 | Rural area |  |
| imbank | Do you work bank/agency/locum shifts? |  |  |  |
|  |  | 1 | Yes |  |
|  |  | 2 | No | Skip to imevdis1 |
| imbanknum | If yes, roughly how many bank/agency/locum shifts do you work per month? |  |  |  |
|  |  | 1 | 1 |  |
|  |  | 2 | 2-5 |  |
|  |  | 3 | 6+ |  |

SECTION 2) Everyday Discrimination

The following statements relate to experiences you may have in your day-to-day life. Please indicate the frequency with which they occur.

| **Name** | **Question** |  |  |  |  |  |
| --- | --- | --- | --- | --- | --- | --- |
|  |  | Very often (1) | Fairly often (2) | Sometimes (3) | Almost never (4) | Never (5) |
| Imevdis1 | You are treated with less courtesy than other people (1) |  |  |  |  |  |
| Imevdis2 | You are treated with less respect than other people (2) |  |  |  |  |  |
| Imevdis3 | You receive poorer service than other people at restaurants or stores (3) |  |  |  |  |  |
| Imevdis4 | People act as if they think you are not smart (4) |  |  |  |  |  |
| Imevdis5 | People act as if they are afraid of you (5) |  |  |  |  |  |
| Imevdis6 | People act as if they think you are dishonest (6) |  |  |  |  |  |
| Imevdis7 | People act as if they are better than you (7) |  |  |  |  |  |
| Imevdis8 | You are called names or insulted (8) |  |  |  |  |  |
| Imevdis9 | You are threatened or harassed (9) |  |  |  |  |  |
| Imevdis10 | You are followed around in stores (10) |  |  |  |  |  |

| **Name** | **Question** | **No** | **Option** | **Skip** |
| --- | --- | --- | --- | --- |
| Imevdis11 | What do you think is the main reason for these experiences? |  |  | Skip if imevdis1-imevdis10 are all “never” |
|  |  | 1 | Ethnic background |  |
|  |  | 2 | Race (skin colour) |  |
|  |  | 3 | Gender |  |
|  |  | 4 | Religion |  |
|  |  | 5 | Sexual orientation |  |
|  |  | 6 | Disability |  |
|  |  | 7 | Age |  |
|  |  | 8 | Weight |  |
|  |  | 9 | Education, income or social class |  |
|  |  | 10 | Mental health problems |  |
|  |  | 11 | Don’t know |  |

SECTION 3) Major Discrimination

The following questions are about situations in which people have sometimes experienced unfair treatment. This may have happened for many reasons, including their ancestry or national origins, gender, race/ethnicity, age etc. Please indicate if any of the following has ever happened to you because of unfair treatment.

At any time in your life, have you been…

| **Name** | **Question** | **Select one** | | **What do you think was the main reason for this experience?** |
| --- | --- | --- | --- | --- |
| Immajdiscrim1 | Unfairly fired? | Yes (1) | No (2) | Ethnic background (1) |
| Immajdiscrim2 | Unfairly not hired for a job? |  |  | Race (skin colour) (2) |
| Immajdiscrim3 | Unfairly denied a promotion? |  |  | Gender (3) |
| Immajdiscrim4 | Unfairly stopped, searcher, questioned, physically threatened or abused by the police? |  |  | Religion (4) |
| Immajdiscrim5 | Unfairly discouraged from continuing your school education by a teacher? |  |  | Sexual orientation (5) |
| Immajdiscrim6 | Unfairly discouraged from continuing your healthcare training? |  |  | Disability (6) |
| Immajdiscrim7 | Unfairly prevented from buying or renting a house/flat by a landlord or estate/leasing agent? |  |  | Age (7) |
| Immajdiscrim8 | Unfairly treated when getting physical healthcare? |  |  | Weight (8) |
| Immajdiscrim9 | Unfairly treated when getting mental healthcare? |  |  | Education, income or class (9) |
|  |  |  |  | Mental health problems (10) |
|  |  |  |  | Don’t know (11) |

| **Name** | **Question** | **No** | **Option** | **Skip** |
| --- | --- | --- | --- | --- |
| Immajdiscrim10 | Have you been unfairly treated in the workplace due to taking maternity/paternity leave? |  |  |  |
|  |  | 1 | Yes |  |
|  |  | 2 | No |  |
|  |  | 3 | Not applicable |  |

SECTION 4) Workplace Discrimination and Harassment

The next set of questions relate to your personal **experiences** of harassment, bullying or abuse at work. This refers to behaviour that is repeated and intended to hurt someone physically or emotionally.

*We understand that acts of discrimination and harassment can occur for multiple reasons. We kindly ask that for the next set of questions you pick the one reason which best captures your experience as this will allow us to better analyse your data. We aim to understand more about the complex reasons for discrimination and harassment in the NHS through our interview study (please visit tidesstudy.com to learn more about this).*

In the last 12 months, how many times have you personally EXPERIENCED harassment, bullying or abuse from…

| **Name** | **Question** | **How many times?** | **What do you think was the main reason for this experience?** |
| --- | --- | --- | --- |
| Imworkexphba1 | Patients/service users, their relatives, or other members of the public? | Never (1) | Ethnic background (1) |
| Imworkexphba2 | Colleagues? | 1-2 (2) | Race (skin colour) (2) |
| Imworkexphba3 | Managers? | 3-5 (3) | Gender (3) |
|  |  | 6-10 (4) | Religion (4) |
|  |  | 10+ (5) | Sexual orientation (5) |
|  |  |  | Disability (6) |
|  |  |  | Age (7) |
|  |  |  | Weight (8) |
|  |  |  | Education, income or class (9) |
|  |  |  | Mental health problems (10) |
|  |  |  | Don’t know (11) |

The next set of questions relate to your personal **experiences** of discrimination (unequal treatment of persons or groups) at work.

In the last 12 months have your personally EXPERIENCED discrimination at work from any of the following?

| **Name** | **Question** | **Experienced** | **What do you think was the main reason for this experience?** |
| --- | --- | --- | --- |
| Imworkexpdisc1 | Patients/service users, their relatives, or other members of the public? | Yes (1) | Ethnic background (1) |
| Imworkexpdisc2 | Colleagues? | No (2) | Race (skin colour) (2) |
| Imworkexpdisc3 | Managers? |  | Gender (3) |
|  |  |  | Religion (4) |
|  |  |  | Sexual orientation (5) |
|  |  |  | Disability (6) |
|  |  |  | Age (7) |
|  |  |  | Weight (8) |
|  |  |  | Education, income or class (9) |
|  |  |  | Mental health problems (10) |
|  |  |  | Don’t know (11) |

The next set of questions relate to your **witnessing** harassment, bullying or abuse of others at work. This refers to behaviour that is repeated and intended to hurt someone physically or emotionally.

In the last 12 months, how many times have you personally WITNESSED harassment, bullying or abuse from…

| **Name** | **Question** | **How many times?** | **What do you think was the main reason for this experience?** |
| --- | --- | --- | --- |
| Imworkwithba1 | Patients/service users, their relatives, or other members of the public? | Never (1) | Ethnic background (1) |
| Imworkwithba2 | Colleagues? | 1-2 (2) | Race (skin colour) (2) |
| Imworkwithba3 | Managers? | 3-5 (3) | Gender (3) |
|  |  | 6-10 (4) | Religion (4) |
|  |  | 10+ (5) | Sexual orientation (5) |
|  |  |  | Disability (6) |
|  |  |  | Age (7) |
|  |  |  | Weight (8) |
|  |  |  | Education, income or class (9) |
|  |  |  | Mental health problems (10) |
|  |  |  | Don’t know (11) |

The next set of questions relate to your **witnessing** discrimination (unequal treatment of persons or groups) of others at work.

In the last 12 months have you WITNESSED discrimination at work from any of the following?

| **Name** | **Question** | **Experienced** | **What do you think was the main reason for this experience?** |
| --- | --- | --- | --- |
| Imworkwitdisc1 | Patients/service users, their relatives, or other members of the public? | Yes (1) | Ethnic background (1) |
| Imworkwitdisc2 | Colleagues? | No (2) | Race (skin colour) (2) |
| Imworkwitdisc3 | Managers? |  | Gender (3) |
|  |  |  | Religion (4) |
|  |  |  | Sexual orientation (5) |
|  |  |  | Disability (6) |
|  |  |  | Age (7) |
|  |  |  | Weight (8) |
|  |  |  | Education, income or class (9) |
|  |  |  | Mental health problems (10) |
|  |  |  | Don’t know (11) |

SECTION 5) Anticipated Discrimination

For the next set of questions, please indicate whether you have stopped yourself from doing certain things because you thought you **might** experience unfair treatment. Have you ever stopped yourself…

| **Name** | **Question** | **Select one** | | **What do you think was the main reason for this experience?** |
| --- | --- | --- | --- | --- |
| Imantdiscrim1 | Applying for a job? | Yes (1) | No (2) | Ethnic background (1) |
| Imantdiscrim2 | Applying for training/education? |  |  | Race (skin colour) (2) |
| Imantdiscrim3 | Applying for a promotion? |  |  | Gender (3) |
| Imantdiscrim4 | Contacting health services for a health complaint? |  |  | Religion (4) |
|  |  |  |  | Sexual orientation (5) |
|  |  |  |  | Disability (6) |
|  |  |  |  | Age (7) |
|  |  |  |  | Weight (8) |
|  |  |  |  | Education, income or class (9) |
|  |  |  |  | Mental health problems (10) |
|  |  |  |  | Don’t know (11) |

SECTION 6) Long-standing Illness

The following questions relate to any long-standing health problems you might have. By long-standing, this means anything that has troubled you over a period of time, or that is likely to affect you over a period of time in the future. This includes recently identified problems.

| **Name** | **Question** | **No** | **Option** | **Skip** |
| --- | --- | --- | --- | --- |
| Imlsi1 | Do you have any long-standing health problems, illness or disability? |  |  |  |
|  |  | 1 | Yes |  |
|  |  | 2 | No | Skip to imlsi3 |
| Imlsi2 | Please specify which long-standing health problems you have. Select all that apply. |  |  |  |
|  |  | 1 | Asthma |  |
|  |  | 2 | Chronic bronchitis |  |
|  |  | 3 | Depression or other nervous illness |  |
|  |  | 4 | Stomach or other digestive disorder |  |
|  |  | 5 | Liver disease |  |
|  |  | 6 | Rheumatic disorder or arthritis |  |
|  |  | 7 | Heart problem |  |
|  |  | 8 | HIV |  |
|  |  | 9 | Stroke |  |
|  |  | 10 | High blood pressure |  |
|  |  | 11 | Migraine |  |
|  |  | 12 | Epilepsy |  |
|  |  | 13 | Gynaecological problems |  |
|  |  | 14 | Cancer |  |
|  |  | 15 | Obesity |  |
|  |  | 16 | Mental health condition |  |
|  |  | 17 | Other |  |
| Imlsi3 | Does your health in any way limit your daily activities compared to most people of your age? |  |  |  |
|  |  | 1 | Yes |  |
|  |  | 2 | No |  |
| Imlsi4 | Does your health limit the type of work or the amount of work you can do? |  |  |  |
|  |  | 1 | Yes |  |
|  |  | 2 | No |  |

SECTION 7) Mental health and wellbeing – PHQ9

The next questions are in reference to your mental health. Over the last 2 weeks, how often have you been bothered by any of the following problems?

| **Name** | **Question** |  |  |  |  |
| --- | --- | --- | --- | --- | --- |
|  |  | Not at all (1) | Several days (2) | More than half the days (3) | Nearly every day (4) |
| Imphq9_1 | Little interest or pleasure in doing things (1) |  |  |  |  |
| Imphq9_2 | Feeling down, depressed or hopeless (2) |  |  |  |  |
| Imphq9_3 | Trouble falling asleep, staying asleep, or sleeping too much (3) |  |  |  |  |
| Imphq9_4 | Feeling tired or having little energy (4) |  |  |  |  |
| Imphq9_5 | Poor appetite or overeating (5) |  |  |  |  |
| Imphq9_6 | Feeling bad about yourself - or that you're a failure or have let yourself or your family down (6) |  |  |  |  |
| Imphq9_7 | Trouble concentrating on things, such as reading the newspaper or watching television (7) |  |  |  |  |
| Imphq9_8 | Moving or speaking so slowly that other people have noticed. Or the opposite – being so fidgety and restless that you have been moving around a lot more than usual (8) |  |  |  |  |
| Imphq9_9 | Thoughts that you would be better off dead or of hurting yourself in some way (9) |  |  |  |  |

*We appreciate that you are willing to share your experiences and feelings with the research team. If you would like emotional support, or completing the survey has caused distress, we encourage you to reach out to someone you trust. If you feel that you are in any immediate danger, please call 111. Information on support services available for healthcare practitioners and community members is available on the* [*TIDES website*](https://tidesstudy.com/signposting-booklet/)*.*

SECTION 8) Mental health and wellbeing – GAD7

Over the last 2 weeks, how often have you been bothered by any of the following problems?

| **Name** | **Question** |  |  |  |  |
| --- | --- | --- | --- | --- | --- |
|  |  | Not at all (1) | Several days (2) | More than half the days (3) | Nearly every day (4) |
| Imgad7_1 | Feeling nervous, anxious or on edge? (1) |  |  |  |  |
| Imgad7_2 | Not being able to stop or control worrying? (2) |  |  |  |  |
| Imgad7_3 | Worrying too much about different things? (3) |  |  |  |  |
| Imgad7_4 | Trouble relaxing? (4) |  |  |  |  |
| Imgad7_5 | Being so restless that it is hard to sit still? (5) |  |  |  |  |
| Imgad7_6 | Becoming easily annoyed or irritable? (6) |  |  |  |  |
| Imgad7_7 | Feeling afraid as if something awful might happen? (7) |  |  |  |  |

SECTION 9) Somatic symptoms - PHQ15

The following questions are in reference to your physical health. During the past 4 weeks (28 days), how much have you been bothered by any of the following problems?

| **Name** | **Question** |  |  |  |
| --- | --- | --- | --- | --- |
|  |  | Not bothered at all (1) | Bothered a little (2) | Bothered a lot (3) |
| Imphq15_1 | Stomach pain (1) |  |  |  |
| Imphq15_2 | Back pain (2) |  |  |  |
| Imphq15_3 | Pain in your arms, legs, or joints (knees, hips, elbows, etc.) (3) |  |  |  |
| Imphq15_4 | Menstrual cramps or other problems with your periods (select "Not bothered at all" if N/A) (4) |  |  |  |
| Imphq15_5 | Headaches (5) |  |  |  |
| Imphq15_6 | Chest pain (6) |  |  |  |
| Imphq15_7 | Dizziness (7) |  |  |  |
| Imphq15_8 | Fainting spells (8) |  |  |  |
| Imphq15_9 | Feeling your heart pound or race (9) |  |  |  |
| Imphq15_10 | Shortness of breath (10) |  |  |  |
| Imphq15_11 | Pain or problems during sexual intercourse (11) |  |  |  |
| Imphq15_12 | Constipation, loose bowels, or diarrhoea (12) |  |  |  |
| Imphq15_13 | Nausea, gas or indigestion (13) |  |  |  |
| Imphq15_14 | Feeling tired or having low energy (14) |  |  |  |
| Imphq15_15 | Trouble sleeping (15) |  |  |  |

SECTION 10) Workplace Support

| **Name** | **Question** | **No** | **Option** | **Skip** |
| --- | --- | --- | --- | --- |
| Imwpsupp1 | In general, do you feel supported by your manager in relation to career development and progression? |  |  |  |
|  |  | 1 | Yes |  |
|  |  | 2 | No |  |
| Imwpsupp2 | Do the managers in your organisation treat all people equally regarding career progression? |  |  |  |
|  |  | 1 | Yes |  |
|  |  | 2 | No |  |
| Imwpsupp3 | Since the pandemic began, have you faced any disciplinary action? |  |  |  |
|  |  | 1 | Yes |  |
|  |  | 2 | No |  |

SECTION 11) Work Environment

| **Name** | **Question** | **No** | **Option** | **Skip** |
| --- | --- | --- | --- | --- |
| imwpenv | Do you feel your organisation values you and what you do? |  |  |  |
|  |  | 1 | Always |  |
|  |  | 2 | Most of the time |  |
|  |  | 3 | About half the time |  |
|  |  | 4 | Sometimes |  |
|  |  | 5 | Never |  |
| Imwpenv2 | Do you feel that your opinions are respected by your colleagues? |  |  |  |
|  |  | 1 | Always |  |
|  |  | 2 | Most of the time |  |
|  |  | 3 | About half the time |  |
|  |  | 4 | Sometimes |  |
|  |  | 5 | Never |  |
| Imwpenv3 | Do you feel able to speak out about workplace practices and procedures that you disagree with or have concerns about? |  |  |  |
|  |  | 1 | Always |  |
|  |  | 2 | Most of the time |  |
|  |  | 3 | About half the time |  |
|  |  | 4 | Sometimes |  |
|  |  | 5 | Never |  |
| Imwpenv4 | Which of the following are barriers to speaking out for you? Select all that apply |  |  |  |
|  |  | 1 | Not being taken seriously |  |
|  |  | 2 | Fear of negative impact on work (e.g. affecting career progression or allocation of shifts) |  |
|  |  | 3 | Fear of experiencing negative responses or backlash from colleagues or managers |  |
|  |  | 4 | Previous negative experiences of speaking out |  |
|  |  | 5 | Witnessing negative experiences when others have spoken out |  |
|  |  | 6 | Other |  |
|  |  | 7 | None / NA |  |

During the pandemic have you ever:

| **Name** | **Question** |  |  | **skip** |
| --- | --- | --- | --- | --- |
|  |  | Yes | No |  |
| Imwpethic_1 | Come into work or worked from home despite being or feeling ill in any way? (1) |  |  |  |
| Imwpethic_2 | Felt a sense of duty to go into work and support your colleagues? (2) |  |  |  |
| Imwpethic_3 | Felt the need to prove yourself to other members of staff? (3) |  |  |  |
| Imwpethic_4 | Felt pressure to work harder than your colleagues? (4) |  |  |  |
| Imwpethic_5 | Felt any guilt when self-isolating/quarantining due to exposure (5) |  |  |  |

SECTION 12) Employment, income and illness

Since the pandemic began, have you experienced any of the following:

| **Name** | **Question** |  |  |
| --- | --- | --- | --- |
|  |  | Yes (1) | No (2) |
| Imempillinc_1 | Loss of personal income (1) |  |  |
| Imempillinc_2 | Loss of family income (2) |  |  |
| Imempillinc_3 | Problems managing your finances (3) |  |  |
| Imempillinc_4 | Reduced employment (4) |  |  |
| Imempillinc_5 | Loss of employment (5) |  |  |
| Imempillinc_6 | Reduced ability to access food (6) |  |  |
| Imempillinc_7 | Illness of a family member due to COVID-19 (7) |  |  |
| Imempillinc_8 | Bereavement due to COVID-19 (8) |  |  |

SECTION 13) Personal Protective Equipment (PPE)

| **Name** | **Question** | **No** | **Option** | **Skip** |
| --- | --- | --- | --- | --- |
| imppe | Since the pandemic have you ever been unable to access PPE when at work? |  |  |  |
|  |  | 1 | Yes |  |
|  |  | 2 | No | skip to imppe3 |
| Imppe2 | How often has this happened? |  |  |  |
|  |  | 1 | Once |  |
|  |  | 2 | Twice |  |
|  |  | 3 | Three or more times |  |
| Imppe3 | Have you ever experienced delays in receiving PPE since the pandemic began? |  |  |  |
|  |  | 1 | Yes |  |
|  |  | 2 | No | skip to imppe5 |
| Imppe4 | When did this happen? |  |  |  |
|  |  | 1 | First wave |  |
|  |  | 2 | Second wave |  |
|  |  | 3 | Both waves |  |
| Imppe5 | Did any items of PPE provided not fit you properly? |  |  |  |
|  |  | 1 | Yes |  |
|  |  | 2 | No | Skip to imppe7 |
| Imppe6 | Which did not fit properly? Select all that apply |  |  |  |
|  |  | 1 | Face mask |  |
|  |  | 2 | Eye protection |  |
|  |  | 3 | Visor face shield |  |
|  |  | 4 | Other |  |
| Imppe7 | Does the PPE provide an adequate level of protection for your duties (e.g., aerosol generating procedures)? |  |  |  |
|  |  | 1 | Yes |  |
|  |  | 2 | No |  |
|  |  | 3 | Unsure/don’t know |  |

SECTION 14) Vaccine

| **Name** | **Question** | **No** | **Option** | **Skip** |
| --- | --- | --- | --- | --- |
| Imvaccine | To what extent do you believe a vaccine can help control the spread of COVID-19 |  |  |  |
|  |  | 1 | Strongly agree |  |
|  |  | 2 | Somewhat agree |  |
|  |  | 3 | Neither agree nor disagree |  |
|  |  | 4 | Somewhat disagree |  |
|  |  | 5 | Strongly disagree |  |
| Imvaccine2a | Have you had a COVID-19 vaccine? |  |  |  |
|  |  | 1 | Yes – 1^st^ dose only | skip to imrisktake1 |
|  |  | 2 | Yes – 1^st^ and 2^nd^ dose | skip to imrisktake1 |
|  |  | 3 | No |  |
| Imvaccine2b | If offered, will you take a COVID-19 vaccine when it first becomes available to you? |  |  |  |
|  |  | 1 | Yes | skip to imrisktake1 |
|  |  | 2 | No |  |
|  |  | 3 | Unsure/undecided |  |
| Imvaccine3 | Why not? |  |  |  |
|  |  | 1 | I believe it won’t be effective | Skip to imrisktake1 |
|  |  | 2 | I have concerns about potential negative side effects for me | Skip to imrisktake1 |
|  |  | 3 | I have concerns about the safety of the vaccine | Skip to imrisktake1 |
|  |  | 4 | I don’t believe there is a COVID-19 virus | Skip to imrisktake1 |
|  |  | 5 | Other |  |
| Imvaccine4 | If Other, please specify |  |  |  |
| Imrisktake1 | Have you chosen not to adhere with social/physical distancing measures at any time (e.g., not wearing a mask in areas that require you to do so)? |  |  |  |
|  |  | 1 | Yes |  |
|  |  | 2 | No |  |
| Imrisktake2 | Have you witnessed colleagues not adhering to guidelines for social/physical distancing measures (e.g., not wearing a mask in areas that require you to do so)? |  |  |  |
|  |  | 1 | Yes |  |
|  |  | 2 | No |  |

SECTION 15) Risk Assessments

| **Name** | **Question** | **No** | **Option** | **Skip** |
| --- | --- | --- | --- | --- |
| imrisk | Have you had a work based COVID-19 risk assessment? |  |  |  |
|  |  | 1 | Yes |  |
|  |  | 2 | No | skip to imredeploy |
| Imrisk2 | Did you find your COVID-19 risk assessment beneficial? (i.e. did it help to manage work activities or situations in a way that reduced your risk of contracting COVID-19 at work?)e |  |  |  |
|  |  | 1 | Yes |  |
|  |  | 2 | No |  |
|  |  | 3 | Unsure |  |
| Imrisk3 | Were any recommendations from your risk assessment(s) followed up by your employers? |  |  |  |
|  |  | 1 | Yes |  |
|  |  | 2 | No |  |
| Imrisk4 | Were you given copies of your risk assessments? |  |  |  |
|  |  | 1 | Yes |  |
|  |  | 2 | No |  |
| Imrisk5 | Did you have a say in decisions made as a result of your risk assessment(s)? |  |  |  |
|  |  | 1 | Yes |  |
|  |  | 2 | No |  |

SECTION 16) Redeployment

| **Name** | **Question** | **No** | **Option** | **Skip** |
| --- | --- | --- | --- | --- |
| imredeploy | At any time during the pandemic, were you redeployed into different areas/roles? |  |  |  |
|  |  | 1 | Yes |  |
|  |  | 2 | No | skip to imredeploy6 |
| Imredeploy2 | Were you forewarned about this redeployment? |  |  |  |
|  |  | 1 | Yes |  |
|  |  | 2 | No |  |
| Imredeploy3 | Did you have any input into the decision about your redeployment? |  |  |  |
|  |  | 1 | Yes |  |
|  |  | 2 | No |  |
| Imredeploy4 | Did you feel able to challenge this decision if you wanted to? |  |  |  |
|  |  | 1 | Yes |  |
|  |  | 2 | No |  |
| Imredeploy5 | Will you/have you been be able to return to your original position following redeployment? |  |  |  |
|  |  | 1 | Yes |  |
|  |  | 2 | No |  |
|  |  | 3 | Unsure |  |
| Imredeploy6 | How confident are you that your opinions/decisions about redeployment would be listened to/acted upon? |  |  |  |
|  |  | 1 | Extremely confident |  |
|  |  | 2 | Very confident |  |
|  |  | 3 | Moderately confident |  |
|  |  | 4 | Slightly confident |  |
|  |  | 5 | Not at all confident |  |
| Imredeploy7 | Do you feel that you have a good understanding of your employment rights in relation to redeployment? (e.g., pay protection) |  |  |  |
|  |  | 1 | Yes |  |
|  |  | 2 | No |  |

SECTION 17) Home-working

| **Name** | **Question** | **No** | **Option** | **Skip** |
| --- | --- | --- | --- | --- |
| imworkhome | Have you been working from home during the pandemic? |  |  |  |
|  |  | 1 | Yes |  |
|  |  | 2 | No – not allowed | skip to imhhldcomp |
|  |  | 3 | No – did not want to | skip to imhhldcomp |
| Imworkhome2 | Did you work from home before the pandemic? |  |  |  |
|  |  | 1 | Yes |  |
|  |  | 2 | No – not allowed |  |
|  |  | 3 | No – did not want to |  |
| Imworkhome3 | Since the pandemic, how often do you work from home? |  |  |  |
|  |  | 1 | Less than once a week |  |
|  |  | 2 | Once a week |  |
|  |  | 3 | 2-3 times a week |  |
|  |  | 4 | 4-6 times a week |  |
|  |  | 5 | Daily |  |
| Imworkhome4 | Do you have adequate equipment to enable you to work effectively from home? |  |  |  |
|  |  | 1 | Yes |  |
|  |  | 2 | No |  |
| Imworkhome5 | Do you have fast/reliable internet access to work from home? |  |  |  |
|  |  | 1 | Yes |  |
|  |  | 2 | No |  |
| Imworkhome6 | Did your organisation provide you with the equipment/secure network access required to work from home? |  |  |  |
|  |  | 1 | Yes |  |
|  |  | 2 | No – not provided |  |
|  |  | 3 | No – not required |  |

SECTION 18) Household Composition

| **Name** | **Question** | **No** | **Option** | **Skip** |
| --- | --- | --- | --- | --- |
| imhhldcomp | Who do you live with? Select all that apply |  |  |  |
|  |  | 1 | Alone | Skip to imshield |
|  |  | 2 | With partner/spouse | Skip to imshield |
|  |  | 3 | With flatmates | Skip to imshield |
|  |  | 4 | With family |  |
| Imhhldcomp2 | Which family members do you live with? Select all that apply |  |  |  |
|  |  | 1 | Child aged under 16 |  |
|  |  | 2 | Child aged 16 or over |  |
|  |  | 3 | Parents |  |
|  |  | 4 | Grandparents |  |
|  |  | 5 | Other |  |
| imshield | Do you currently live with someone who is/was shielding? |  |  |  |
|  |  | 1 | Yes |  |
|  |  | 2 | No |  |
| imcaregive | Do you provide unpaid care to a family member, partner or friend who needs help because of their illness, frailty, disability, physical or mental health problem, or an addiction? |  |  |  |
|  |  | 1 | Yes |  |
|  |  | 2 | No | Skip to end |
| Imcaregive2 | Who is this for? Select all that apply |  |  |  |
|  |  | 1 | Child aged under 16 |  |
|  |  | 2 | Child aged 16 or older |  |
|  |  | 3 | Parent |  |
|  |  | 4 | Partner |  |
|  |  | 5 | Other family |  |
|  |  | 6 | Other |  |

We thank you for your time spent taking this survey. Your response has been recorded.  If you would like to know more about the TIDES study including free [workshops and project updates](https://tidesstudy.com/microaggressions-workshop/), please visit our website tidesstudy.com or email at . Please feel free to leave any comments about any of the topics raised in this survey at tidesstudy.com/have-your-say

### Supplementary Material C: Census grouping of ethnicities

**Asian or Asian British**

- Indian
- Pakistani
- Bangladeshi
- Chinese
- Any other Asian background

**Black, Black British, Caribbean, or African**

- Caribbean
- African
- Any other Black, Black British, or Caribbean background

**Mixed or multiple ethnic groups**

- White and Black Caribbean
- White and Black African
- White and Asian
- Any other Mixed or multiple ethnic background

**White**

- English, Welsh, Scottish, Northern Irish or British
- Irish
- Gypsy or Irish Traveller
- Roma
- Any other White background

**Other ethnic group**

- Arab
- Any other ethnic group

### Supplementary Material D: Subgroup analyses

**Table D1: Subgroup analysis of probable depression by ethnicity, stratified by BHA and discrimination**

|  | **No BHA** | **BHA** | **No Discrimination** | **Discrimination** |
| --- | --- | --- | --- | --- |
|  | n (%) | n (%) | n (%) | n (%) |
| **Probable depressive disorder** |  |  |  |  |
| White British | 350 (15.6) | 337 (34.4) | 467 (17.4) | 220 (42.9) |
| White Other | 35 (18.0) | 48 (39.2) | 50 (19.3) | 33 (44.5) |
| Black | 14 (21.7) | 15 (25.6) | 13 (18.1) | 16 (30.1) |
| Asian | 22 (14.1) | 25 (37.1) | 23 (13.8) | 24 (45.2) |
| Mixed/Other | 12 (22.8) | 15 (46.8) | 12 (19.2) | 15 (55.7) |
| BHA = Bullying, Harassment or Abuse | | | | |
| Probable depressive disorder = PHQ-9 score of ≥10 | | | | |

### Supplementary Material E: List of participating NHS Trusts

The following NHS Trust participated in the NHSCHECK study.

**Trust names**

Avon & Wiltshire Mental Health Partnership NHS Trust

Cambridge University Hospitals NHS Foundation Trust

Cambridgeshire and Peterborough NHS Trust

Cornwall Partnership Trust

Devon Partnership NHS Trust

East Suffolk and North Essex NHS Trust

Gloucestershire Hospitals NHS Trust

Guy's and St Thomas' NHS Trust

King's College Hospital and PRUH

Lancashire and South Cumbria NHS Trust

Norfolk and Norwich University Hospitals

Nottinghamshire Healthcare NHS Trust

Royal Papworth Hospital

Sheffield Health and Social Care NHS Foundation Trust

South London and Maudsley (SLAM)

Tees Esk and Wear Valleys NHS Foundation Trust

University Hospitals of Derby and Burton

University Hospitals of Leicester NHS Trust
